# Speech pause distribution as an early marker for Alzheimer’s disease

**DOI:** 10.1101/2020.12.28.20248875

**Authors:** Patricia Pastoriza-Domínguez, Iván G. Torre, Faustino Diéguez-Vide, Isabel Gómez-Ruiz, Sandra Geladó, Joan Bello-López, Asunción Ávila-Rivera, Jordi Matías-Guiu, Vanesa Pytel, Antoni Hernández-Fernández

## Abstract

**Background:** Pause duration analysis is a common feature in the study of discourse in Alzheimer’s disease (AD) and may also be helpful for its early detection. However, studies involving patients with amnestic mild cognitive impairment (aMCI) have yielded varying results.

**Objectives:** To characterize the probability density distribution of speech pause durations in AD, two multi-domain amnestic MCI patients (with memory encoding deficits, a-mdMCI-E, and with retrieval impairment only, a-mdMCI-R) and healthy controls (HC) in order check whether there are significant differences between them.

**Method:** 112 picture-based oral narratives were manually transcribed and annotated for the automatic extraction and analysis of pause durations. Different probability distributions were tested for the fitting of pause durations while truncating shorter ranges. Recent findings in the field of Statistics were considered in order to avoid the inherent methodological uncertainty that this type of analysis entails.

**Results:** A lognormal distribution (LND) explained the distribution of pause duration for all groups. Its fitted parameters (*µ,σ*) followed a gradation from the group with shorter durations and a higher tendency to produce short pauses (HC) to the group with longer pause durations and a considerably higher tendency to produce long pauses with greater variance (AD). Importantly, a-mdMCI-E produced significantly longer pauses and with greater variability than their a-mdMCI-R counterparts (*α* = 0.05).

**Conclusion:** We report significant differences at the group level in pause distribution across all groups of study that could be used in future diagnostic tools and discuss the clinical implications of these findings, particularly regarding the characterization of aMCI.

## Introduction

With nearly half of the individuals diagnosed with Mild Cognitive Impairment (MCI) due to Alzheimer’s Disease (AD) developing dementia within three years [1,2], it has been suggested that diagnosis at the prodromal stage represents the optimal time-window for onset delay and potential intervention [3]. Not only is it more cost-effective than diagnosis at the dementia stage [4], but it is also considered to be less distressful for patients in comparison to population screenings, allowing for care planning when concerns are raised and assistance is needed [5]. The assessment of the constellation of cognitive deficits found in the prodromal stage of AD has proven to provide considerable diagnostic power regarding dementia progression regardless of biomarker use [6] and seems to have clearer clinical utility [2,7]. Crucially, neuropsychological testing is accessible in every clinical context, which makes cognitive profiling the current focus of general clinical practice [9] and prevention efforts [10]. However, due to the heterogeneity in MCI’s clinical profile and outcome, risk factors for Alzheimer’s dementia and their association with MCI subtypes need yet to be ascertained [11].

While the prognostic relevance of the number of cognitive domains affected is still under debate [11,1,12], the presence of memory deficits –with or without accompanying impairment in other domains– was initially considered a definitory feature of the clinical syndrome [13,14,15] and is a consistent factor in the clinical profile of a great proportion of MCI patients with a later progression to dementia [16,17,18,19,20,21]. More specifically, impairment in episodic memory encoding reflected in unsuccessful cued recall and susceptibility to intrusion effects has been associated with higher probability of underlying [22,23,24] even at the prodromal phase of MCI [25]. More recent studies have demonstrated that this particular cognitive profile in MCI is compatible with positive AD biomarkers [26,27,28,29,30], as well as abnormal neural connectivity [31,32,33] and cerebral perfusion patterns [34] during memory encoding tasks. Moreover, declines in memory encoding have been predicted in amyloid-positive cognitively-normal individuals [35,36,37] alongside downstream tau increases [38,39,40,41], as well as in patients with subjective cognitive complaint with positive AD biomarkers [42]. Abnormal cortical activity has also been observed in genetic carriers during memory encoding tasks [43,44], supporting the thesis of a preference of AD pathology for cortical areas devoted to memory processing and, more precisely, those involved in encoding and retrieval of newly learned information.

Language impairment has also been consistently observed in MCI patients who end up with a dementia diagnosis [45,46]. AD progressors perform significantly worse than non-progressors at naming [47,48,50,49] and semantic fluency [50,51,52,53] with prediction models combining biomarkers and composite cognitive scores benefitting significantly from the inclusion of language-related scores[16,54,55]. Spontaneous speech is a highly complex process recruiting various levels of linguistic processing that has revealed substancial differences between patients with MCI due to Alzheimer’s Disease (AD) and their healthy counterparts, predominantly in the form semantic impoverishment and reduced fluency (see [56,57] for recent reviews).

The great advances in fields such as natural language processing and automatic speech recognition in the last decades have contributed to a surge in studies reporting in most cases on a large number of linguistic and paralinguistic variables [58,59,60,61,62,63,64,65,66,67,68,69], ranging from voice features [70,71,72] to discourse analysis [73,74,19,75,76] in the characterization of patients with MCI due to AD. The measurement of voiced and unvoiced segments in connected speech has been a particularly prolific research avenue thanks to its relative methodological simplicity and the great technical precision that current technologies grant. In this regard, it has been found that AD patients produce more silent pauses than healthy controls (HC) [77], more frequently [78,79] – although not in [80]- and with longer mean duration [79,80] (but not in [77]), thus representing a larger proportion of discourse time in comparison to voiced segments [78,79,62,81].

These speech fluency features are already present in MCI, with patients producing more pauses [69] at a higher rate than HC [59,68], although pause rate did not differ in other studies [69,82] and neither did the number of pauses [83]. Mean pause duration is another recurring feature of study since patients with MCI are expected to produce pauses with longer mean duration than those of healthy controls [62,59,82], although in other studies no significant difference was found [68,83] or this was finding was task-dependent [69]. Voice-to-silence ratio seems to be a more reliable feature, consistently differentiating the narratives of patients with MCI from their healthy counterparts with a lower proportion of voiced time in relation to total discourse time [50,69,59,68,83].

Differences in task choice, criteria for pause labelling, temporal thresholding, or methodology applied-manual versus automatic transcription and segmentation-may have contributed to some extent to discrepancies in the results, as seem to point out studies assessing different task types as [69,59] in MCI and [79] in AD or comparing manual and automatic annotation [68].

In previous studies it has been pointed out that speech segments are not normally distributed and that, therefore, moments of the distribution –e.g. mean and variance– may be inadequate for the characterization of linguistic elements including words and speech pauses [84,85,86,87,88,89]. Further studies, have shown that lognormal distribution is a more accurate approximation for the description of speech pause duration distribution in human voice, showing consistency across studies and good sensitivity for identifying particular constraints such as distribution across discursive and syntactic boundaries or task type [90,91,92]. Its application to language-impaired groups in other neurological disorders such as vascular aphasia or ataxic dysarthria has confirmed these findings, revealing differences in duration and distribution between those and healthy controls [86,93,94].

In this work we firstly endeavour the characterization of the distribution of speech pauses in AD, amnestic MCI and HC considering some limitations inherent to the segmentation of speech pauses such as higher relative errors on shorter ranges. For this purpose, we address the fitting of truncated distribution considering recent discussions on cut-off point selection on long-tail distributed data. In particular, we show that patients with confirmed Alzheimer’s dementia and amnestic multi-domain MCI patients with memory encoding deficits (a-mdMCI-E) at high risk of dementia progression produce significantly longer pauses and with more dispersion than healthy controls, as well as to determine the relative weight of long pauses in relation to their general pattern of pause production. Crucially, we expect to also find significant differences in pause duration and distribution between the group with a-mdMCI-E and their a-mdMCI counterparts with retrieval impairment only (a-mdMCI-R), since the former are more prone to AD progression that the latter. We discuss the validity of pause analysis in the prediction of MCI outcome in the AD spectrum, contributing to the refinement of the current clinical profile of MCI due to AD and to the race for non-invasive, low-cost diagnostic tools for dementia diagnosis.

## 1. Materials and methods

### 1.1. Participants

Patients were recruited prospectively and retrospectively through the Neurology units of the Hospital General de Hospitalet de Llobregat (Hospitalet de Llobregat), Hospital Moisés Broggi (Sant Joan Despí) and Hospital Clínico San Carlos (Madrid) during the period 2015 to 2019. Healthy controls included patients’ relatives and volunteers. A total of 112 participants aged 58 to 91 were recruited for the purpose of this study.

Probable AD diagnosis was based on the criteria of the National Institute of Neurological and Communicative Disorders and Stroke/Alzheimer’s Disease and Related Disorders Association (NINCDS-ADRDA) [95] and the National Institute on Aging and Alzheimer’s Association (NIA-AA) [96] for the retrospective and prospective cohort, respectively. All patients included in this group (*n* = 26) had a Clinical Dementia Rating (CDR) [97] score of 1.

56 subjects with MCI were diagnosed according to Petersen criteria [98] in the initial cohort and NIA-AA criteria [99] for prospective participants. More specifically, patients were of the amnestic-multidomain MCI subtype (a-mdMCI) with a CDR score of 0.5. Patients in this group were further classified into two groups according to their memory impairment profile at the Rey Auditory Learning Test (RAVLT) [100]. One subgroup displayed impaired delayed recall but normal recognition memory (deficit in retrieval processes, a-mdMCI-R, 28 participants), and the other one showed both impaired delayed recall and recognition memory (deficit in encoding processes, a-mdMCI-E, 29 participants) [22]. As explained in the introductory section, the latter pattern of impairment has been observed to be more prone to AD progression [23,101]. Follow-up of 16 of the 29 a-mdMCI-E patients and of 18 of the 28 a-mdMCI-R participants revealed that 10 (55%) of the former progressed to AD diagnosis within three years, whereas only one a-mdMCI-R patient (6%) followed the same course.

Additionally, 29 age-matched cognitively unimpaired participants, with no history of neurological disease and a minimum Mini Mental State Examination (MMSE) [101] score of 25 were also recruited as healthy controls (HC). Confirmation of any other neurological condition, history of psychiatric disorder, alcohol abuse or the use of any medication or systemic disease that might justify the observed cognitive impairment was considered a motive for exclusion. None of the participants suffered any visual or hearing impairment that could affect their performance.

### 1.2. Standard protocol approval and patient consent

The study was approved by the Bioethics Committee of Universitat de Barcelona (*IRB00003099*), by the clinical research ethics committees of Hospital Clínico San Carlos (ref. *19/046-E*) and by the Consorci Sanitari Integral-Hospital Universitari de Bellvitge in the case of Hospital General de l’Hospitalet de Llobregat (ref. *19/43-PR222/19*). All participants signed a written informed consent form prior to enrollment in the study.

### 1.3. Neuropsychological protocol

Patient assessment included, in addition to MMSE [101] and the Rey Auditory Verbal Learning Test [100], the 60-item version of the Boston Naming Test (BNT) [102], direct and reverse WAIS digits and Block Design test [103], category (animals) and letter fluency (letter p), the clock-Drawing test [104] and Poppelreuter’s Overlapping Figures Test [105]. Biographic memory was evaluated by means of a five-item questionnaire requesting two important dates-usually a wedding and a relative’s birthdayand the names of three famous people. During the same testing session participants were asked to complete the picture description task of the Bilingual Aphasia Test [106], based on a simple six-picture story depicted on a single sheet of paper – see Figure 1–.

**Figure 1.**
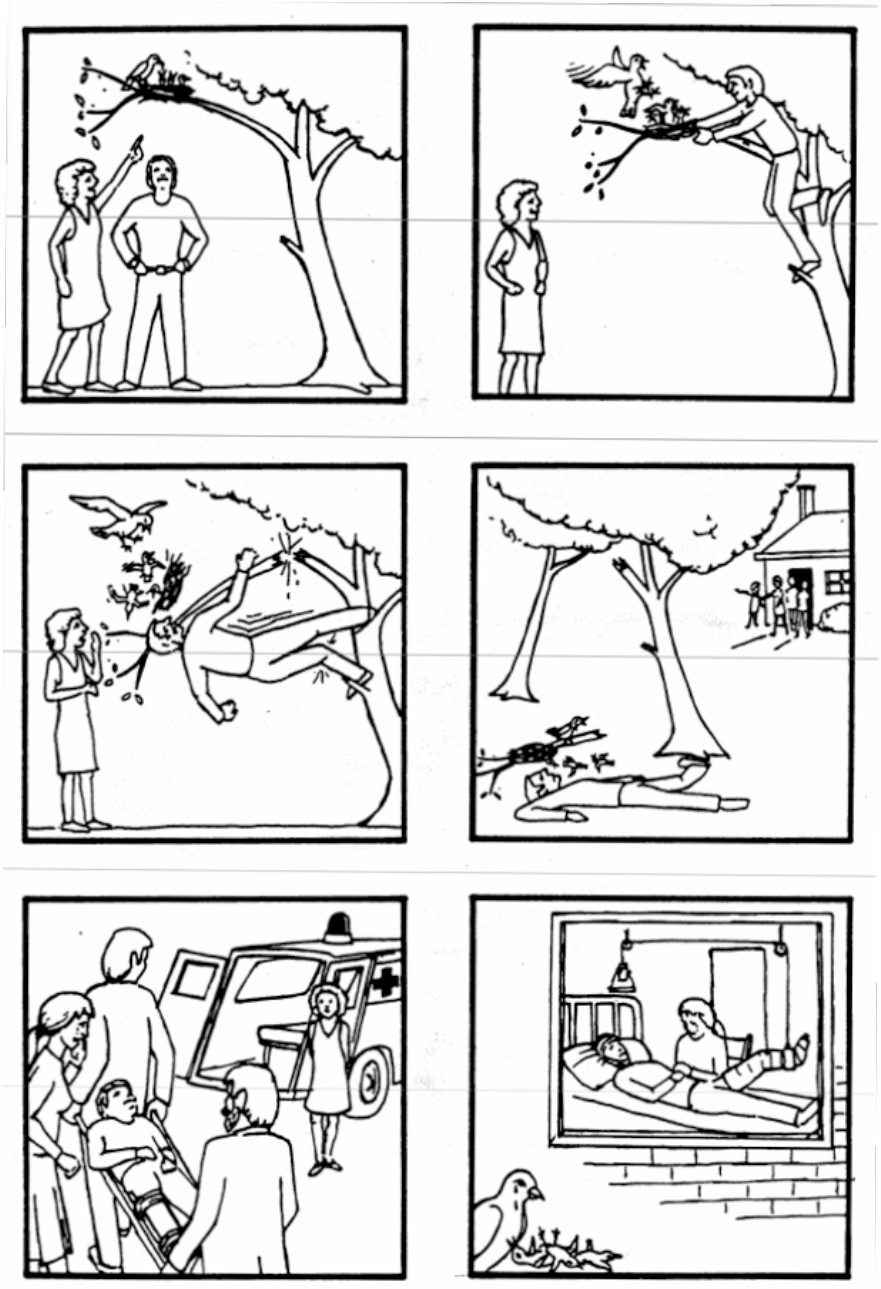
Picture description task. Bilingual Aphasia Test, Paradis (1987) [106].

### 1.4. Participant characteristics

Participant mean age was 76 ± 7 years of age and mean years of education was 6 ± 3 years. Patients in the AD group were significantly older (81 ± 6) than HC (76 ± 8) and both the a-mdMCI-R (75 ± 7) and the a-mdMCI-E (76 ± 5) groups, *F*(3, 111) = 4.58, *p* < 0.05. There were significant differences in years of education across groups (*H*(3) = 16.11, *p* < 0.05) since AD patients (5.6±2.5) and participants in the a-mdMCI-E group (6.1 ± 2.9) had significantly less years of education than HC (8 ± 2). MMSE scores differed significantly (*H*(3) = 64.23, *p* < 10^−3^), with individuals with AD obtaining the lowest mean score (21.8 ± 2.5), which was significantly lower than that of HC (29 ± 1.2), a-mdMCI-R patients (27.4 ± 2.3) and individuals with a-mdMCI-E (25.5 ± 2.6). a-mdMCI-E patients scored significantly lower than HC at MMSE (Bonferroni correction and Dunn post hoc test, *p* < 10^−3^). More detailed information on the demographic characteristics of the sample are provided in Table 1.

**Table 1.**
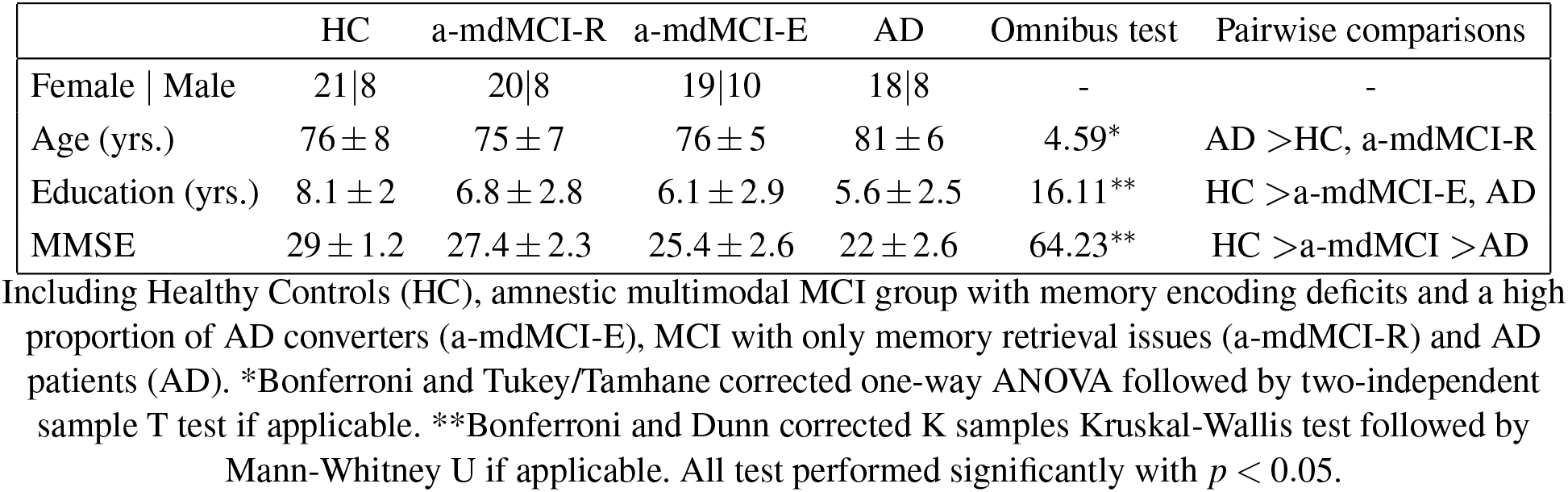
Demographic information.

### 1.5. Neuropsychological profile

Regarding the two a-MCI groups, patients with a-mdMCI-E performed significantly worse than subjects with a-mdMCI-R at MMSE. The only other significant differences between the two groups were observed in their RAVLT performance, with a-mdMCI-E obtaining significantly lower total and delayed recall scores than patients with a-mdMCI-R. No significant differences were found between the two groups in any other cognitive domain as per their scores at BNT, WAIS digit span (direct and reverse), semantic and phonological fluency, WAIS III blocks, Clock Drawing Test, Poppelreuter’s Overlapping Figures Test and remote memory. Patients with AD performed significantly worse than patients in the two a-MCI groups at every test except the two WAIS digits tasks, where no differences were observed amongst the groups of study. Full details and pairwise comparison results are provided in Table 2.

**Table 2.** Neuropsychological profile.

|  | a-mdMCI-R | a-mdMCI-E | AD | Omnibus test | Pairwise comparisons |
| --- | --- | --- | --- | --- | --- |
| RAVLT-total | 26±6.9 | 21.6±6.5 | 15.4±4.9 | 19.31* | a-mdMCI-R >a-mdMCI-E >AD |
| RAVLT-delayed | 3.7±2.2 | 0.6±1.3 | 0.08±0.39 | 51.1** | a-mdMCI-R >a-mdMCI-E, AD |
| WAIS (direct) | 4.4±0.6 | 4.2±0.5 | 4±0.8 | 5.83** | - |
| WAIS (reverse) | 2.7±0.9 | 2.8±0.7 | 2.4±0.6 | 3.63** | - |
| BNT | 48.9±4.6 | 46.3±3.5 | 34.6±5.9 | 67.60* | a-mdMCI-R, a-mdMCI-E >AD |
| Poppelreuter | 10±0 | 10±0 | 9.7±0.5 | 13.99** | a-mdMCI-R, a-mdMCI-E >AD |
| Clock Drawing Test | 3.6±0.8 | 3.6±0.8 | 2±1.3 | 28.62** | a-mdMCI-R, a-mdMCI-E >AD |
| WAIS block design | 20.9±9.6 | 18.5±8.9 | 12.1±6. | 7.04* | a-mdMCI-R, a-mdMCI-E >AD |
| Letter fluency | 8.3±3.8 | 9.2±4.2 | 5.7±3.1 | 11.69** | a-mdMCI-R, a-mdMCI-E >AD |
| Category fluency | 10.1±4.8 | 10.8±4.5 | 6.8±2.6 | 7.3* | a-mdMCI-R, a-mdMCI-E >AD |

\*Bonferroni and Tukey/Tamhane corrected one-way ANOVA followed by two-independent sample T test if applicable. \*\*Bonferroni and Dunn corrected K samples Kruskal-Wallis test followed by Mann-Whitney U if applicable. All test performed significantly with $p < 0.01$ except WAIS (direct) with $p \sim 0.54$ and WAIS (reverse) with $p \sim 0.16$ .

### 1.6. Data acquisition and segmentation

Oral narratives were recorded in a quiet room in hospital by means of a SONY IICD-SX78 recorder at a sampling frequency of 44.1 kHz and subsequently processed in Praat [107] at the same sampling frequency. Audios were transcribed and annotated manually by the first author to allow pause tally and duration extraction by means of a script designed ad hoc.

Pauses were defined as any filled or silent interruption of the speech flow that could not be identified as a linguistic item (such as a disfluency) or as a false start. Filled pauses were thus standardized place-holders that were not lexicalized such as “uhm” or “erm”, as opposing filler expressions such as “bueno” (“well”) or the strategic lengthening of conjuctions, which were labelled as fillers and included in the disfluency tally. While some authors consider these vocalized pauses disfluencies [108,109] or fillers [110,111,112], most studies in the speech and dementia literature count them as filled pauses when explicitly described [69,61,113]. The lower temporal threshold for pause segmentation was set at 50 milliseconds.

### 1.7. Interrater agreement

With the purpose of testing the coherence and replicability of the annotation system 10% of the original 112-narrative corpus (10 narratives) were transcribed and annotated by the fifth author. Comparison of word-by-word transcriptions reached an agreement level of 97.5%, whereas interrater agreement at pause identification was 99.04%. The mean absolute difference of duration measures between the two annotators was 17 *ms* with a Pearson correlation coefficient of 0.99.

### 1.8. Truncated distributions

Following previous work [89], we here consider three possible candidate families of long tail probability density distributions, being all of them defined with two parameters: Lognormal distribution (LND), Gamma distribution and Weibull distribution. LND is known to be generated by multiplicative processes but also additive processes when some conditions are met, as seems recently shown to happen in speech [89]. LNDs are present in many natural systems [114] and have as a property that, being *X* an independent continuous random variable generated by a LND, then the logarithm of *X* is normally distributed. LND, Gamma and Weibull distribution functions are defined as follows:

i. Lognormal distribution:

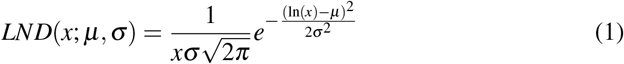
ii. Gamma distribution:

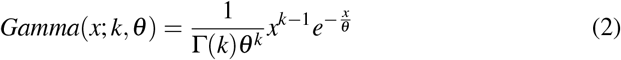

being Γ the gamma function.
iii. Weibull distribution:

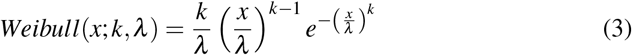

A truncated probability distribution is a distribution whose observations are reduced to some specific range. This technique is particularly useful when it is not possible to have reliable samples for the entire range but the underlying generating dynamic is expected to remain stable, so that the full distribution can be fitted into the truncated observed range. This may be the case for speech segmentation, where shorter samples are subject to higher uncertainty due to factors including Automatic Speech Alignment limitations [115], manual segmentation bias [89] and speaker-driven mixed statistical artifacts (see SI in [89]).

### 1.9. Statistical analysis

Omnibus between-group differences were assessed using one-way ANOVA or Kruskal-Wallis tests as appropriate, followed by pairwise testing with Student T or Mann-Whitney U where applicable.

Lower cut-off point selection when fitting long tail distributions –as those that appears in speech pause duration– is a challenge that has been widely discussed over the last years [116]. In this regard, there has been some agreement on fitting the parameters of the distribution by maximum likelihood estimation (MLE) [117] and using Kolmogorov-Smirnov distance for cut-off point selection [118]. First, for model selection we pick several lower cut-off candidates and fit by MLE the three families of probability density function with the two parameters explained above (Lognormal, Weibull and Gamma). Then Akaike Information Criteria (AIC) and Bayesian Information Criteria (BIC) were used for model selection. Goodness of fit is checked by Kolmogorov Smirnov (KS) testing as to whether reject the distribution or not at the significance level of *p* = 0.05.

Then, we refine the search for the lower cut-off point for the Lognormal distribution by using the method proposed by [119], which is a modified version of [116]. The procedure is as follows. (i) First pick any lower cutoff threshold value, (ii) Fit, by MLE the truncated lognormal distribution to the range *x >*threshold. This lead to the fitted parameters *µ* and *σ*. (iii) Compute Kolmogorov-Smirnov distance *D* between the theoretical distribution with estimated parameters and the real data. (iv) Stochastically generate the same number of samples but from the fitted distribution. (v) Compute Kolmogorov-Smirnov distance *D*_*r*_ between stochastic data and the theoretical distribution. (vi) Repeat 1000 times steps iv and v counting the number of times where *D*_*r*_ < *D*. Repeat this process for different lower cut-off points and select the one where *D*_*r*_ < *D* happens fewer times. Note that [116] showed that there will be a minimum.

## 2. Results

### 2.1. Speech pause duration distribution analysis

We fit pause duration observations from each patient group into three possible theoretical truncated distributions using MLE: Lognormal (LND), Gamma, and Weibull. We use goodness of fit AIC and BIC for model selection (where the lowest the better, see table 3), confirming that, for all cases, pause duration distribution is better explained by a LND. These results are in line with previous reported analysis in speech [89,115].

**Table 3.** LND parameters and goodness of fit.

|  | samples | LND |  | Goodness of fit AIC \| BIC |  |  |
| --- | --- | --- | --- | --- | --- | --- |
| | | $\mu$ | $\sigma$ | LND | Gamma | Weibull |
| HC | 736 | $-0.56 \pm 0.03$ | $0.75 \pm 0.02$ | 779 \| 789 | 792 \| 801 | 794 \| 803 |
| a-mdMCI-R | 679 | $-0.51 \pm 0.03$ | $0.85 \pm 0.03$ | 992 \| 1000 | 979 \| 988 | 979 \| 988 |
| a-mdMCI-E | 618 | $-0.42 \pm 0.04$ | $0.86 \pm 0.03$ | 958 \| 967 | 977 \| 987 | 974 \| 983 |
| AD | 669 | $-0.36 \pm 0.04$ | $0.92 \pm 0.03$ | 1241 \| 1250 | 1272 \| 1281 | 1264 \| 1273 |

(Left) Estimated LND parameters ( $\mu$ , $\sigma$ ) for speech pause duration distributions in each patient group and truncated below 160 *ms*. (Right) AIC and BIC goodness of fit for different alternative distributions (the lower the AIC and BIC the better). All tested distributions are defined by two parameters and within each patient group they are evaluated under the same conditions, so goodness of fit values can be used for model selection.
For almost all cases LND is the most plausible distribution and in all cases LND passes the Kolmogorov Smirnov goodness of fit test at a 95% confidence level.

Lower cut-off point has been calculated following the procedure explained in section 1. Inset panel of Figure 2 shows that there is a minimum on the times that *D*_*r*_ < *D* (*ρ*) at 160 ms which will be chosen as lower threshold. Moreover, main panel of Figure 2 shows the mean relative error between speech pause annotations (blue bars and line) is drastically reduced for speech pauses longer than approximately 150*ms*, while interrater disagreement (1− interrater agreement) is rapidly decreased for pauses longer than 100*ms*. Total number of pauses for each group after the truncation is reflected in table 3, being 736 for HC, 679 for a-mdMCI-R, 618 for a-mdMCI-E and 669 for AD.

**Figure 2.**
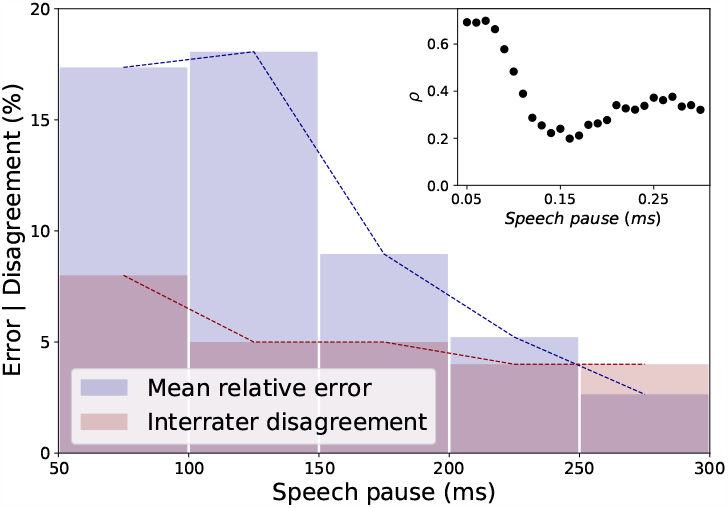
Relative error, interrater disagreement and lower cut-off point estimation. Interrater disagreement (1-interrater agreement) quickly decreases after 100*ms*, while mean relative error of speech pauses that coincide between annotators is quickly reduced after 150*ms*. The inset panel shows cut-off point selection where *ρ* reach a minimum at 160*ms* [116,119].

In addition to this, Kolmogorov Smirnov testing confirms the goodness of fit of the LND at a 95% confidence interval. This can be observed in Figure 3 with the empirical probability of pause duration distribution represented in bars for each group and their fitted truncated LND with solid lines. For the sake of clarity we also provide log-linear representations in the inset panels, where the shape of the LND turns to Gaussian. Estimated LND parameters are listed in table 3, showing that:

**Figure 3.**
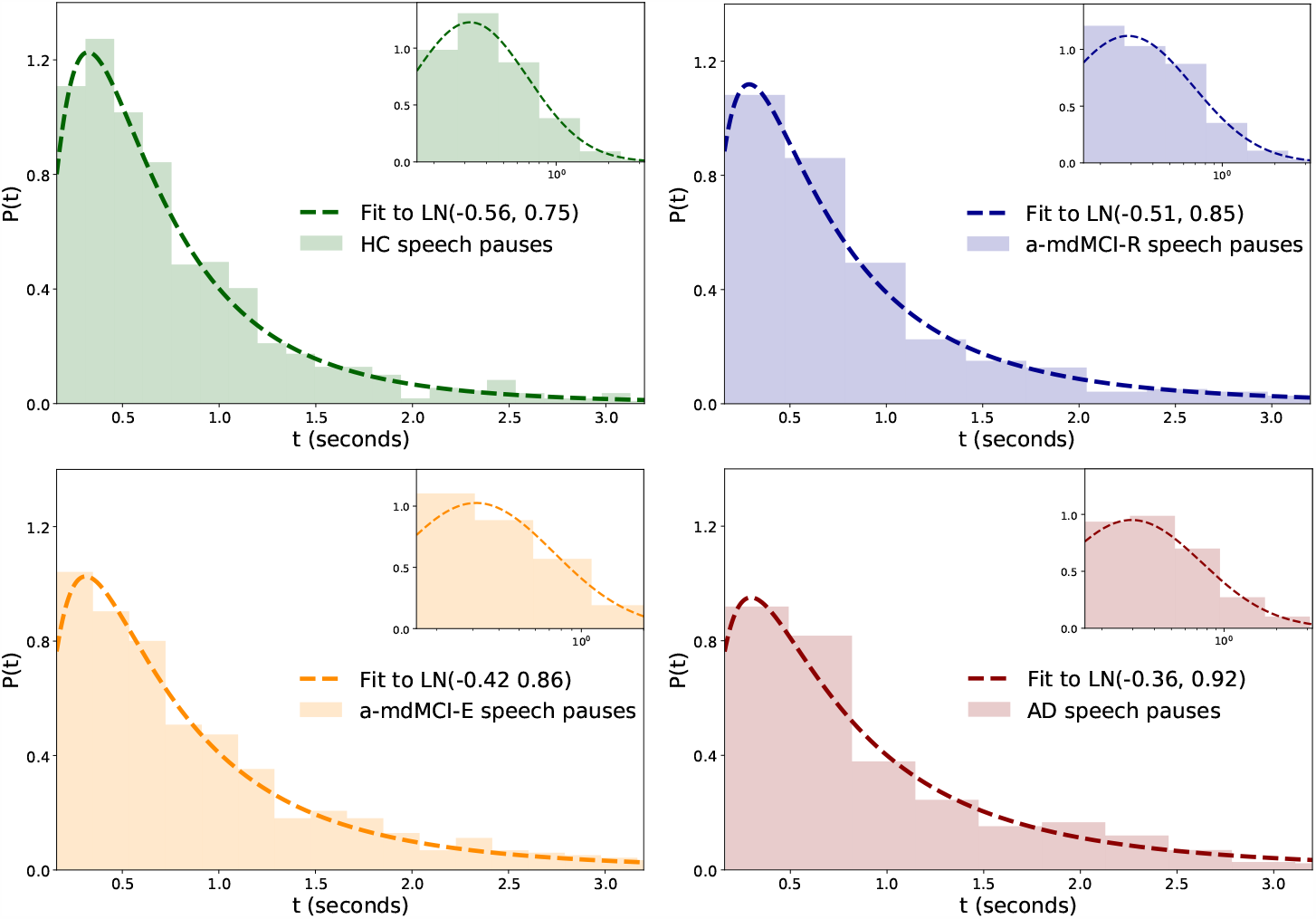
Probability time duration distribution of pauses in each group. For each patient group, the main panel shows in linear axes the probability time duration distribution of pauses –bar representation– and the ML fitted truncated LND. The inner panels are a visual representation of the same results with logarithmic binning and log-linear axis.

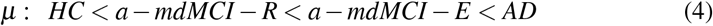

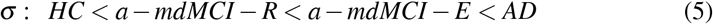

where *µ* represents in LND the multiplicative mean and *σ* is related to more sparse samples, clearly showing that the HC group has a higher probability of making short pauses with a lesser deviation than AD patients (table 3), being mild cognitive impairment groups between them with a-mdMCI-E closer to AD.

Finally, in Figure 4 we represent truncated LNDs with estimated parameters for each group revealing that HC shows higher likelihood of making pauses at range 200*ms* −700*ms* in relation to their total pause production than the AD group, with firstly a-mdMCI-E and subsequently a-mdMCI-R interestingly standing between AD and HCs in the probability gradation. This is just the opposite at longer ranges (*t >* 1.5*s*), where AD patients reveal higher probability than HCs to make long pauses, with the two a-mdMCI groups again performing mid-range and a-mdMCI-E participants displaying a more resembling performance to that of AD patients. Dotted lines represent the continuation of the LNDs out of the truncated range and inner panels express the same results through their log-linear representation.

**Figure 4.**
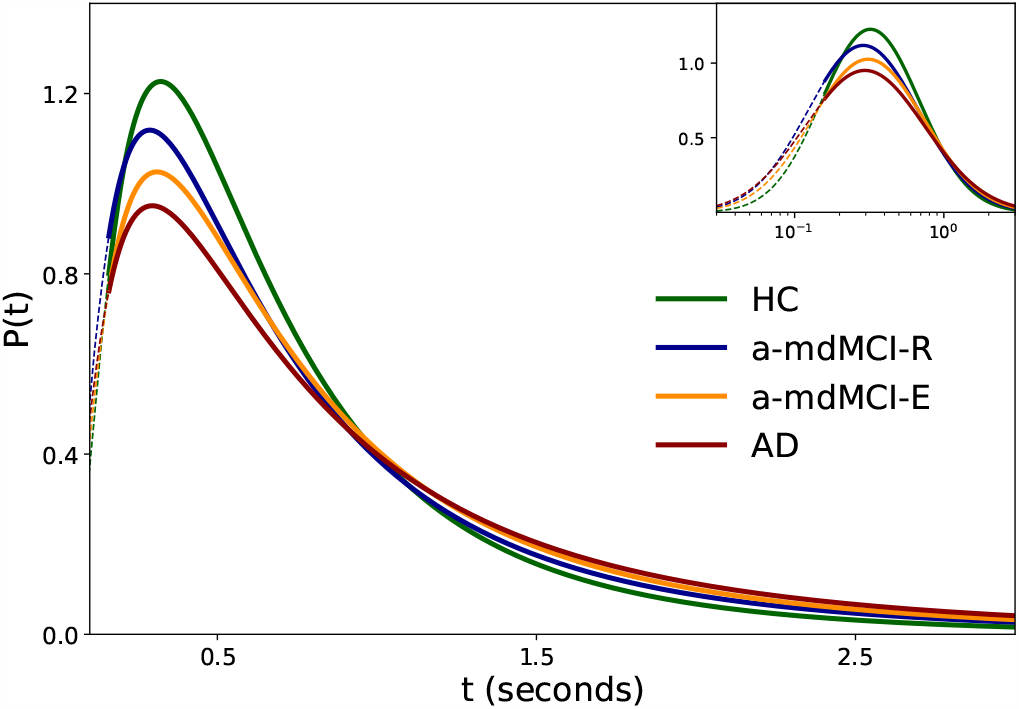
Comparison of time duration distribution between controls and patients groups. The main panel displays the truncated LND-fitted time duration distribution for each group. The HC group shows a higher probability of making short pauses (200-600 *ms*) than the AD group, while AD patients show the opposite pattern with a higher probability of making long pauses (1.5*s* - 2.5 *s*) than HC (tail of the distribution). Interestingly, a-mdMCI-E and a-mdMCI-R always stand in the middle of the continuum for both pause types. Solid lines represent the range with reliable observations, while the dotted line represents the continuation of the LND to shorter timescales. The inner panel shows the same results on the log-linear axis.

Two sided Kolmogorv-Smirnov testing has been used under the null hypothesis that different group samples come from the same distribution and that differences are due to stochastic variations, confirming in all paired cases that differences are significant, therefore rejecting the null hypothesis (see table 5).

**Table 4.**
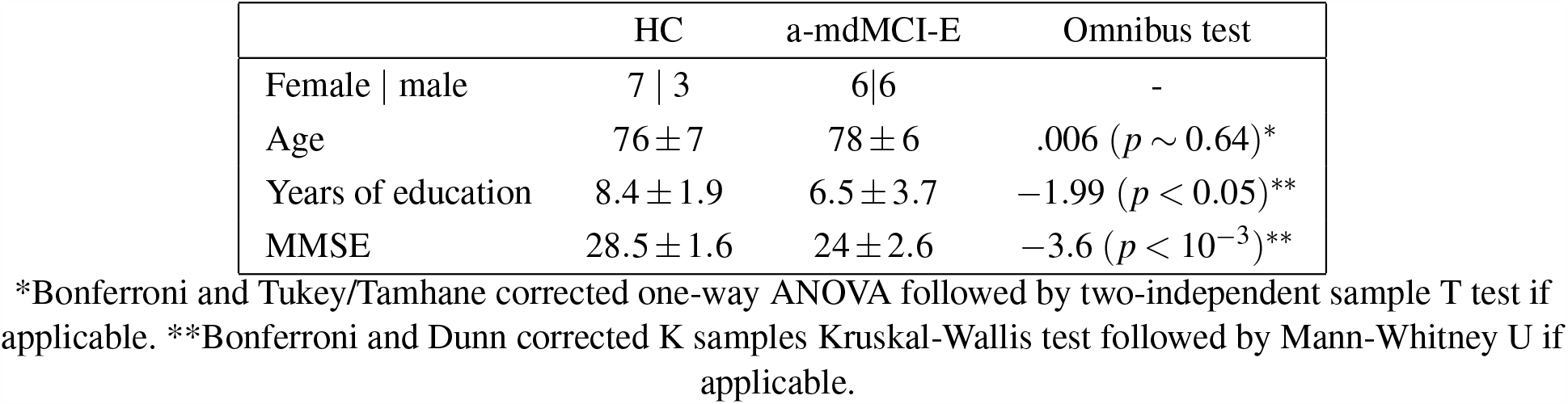
Filtered dataset: demographic information.

|  | HC | a-mdMCI-E | Omnibus test |
| --- | --- | --- | --- |
| Female \| male | 7 \| 3 | 6\|6 | - |
| Age | $76 \pm 7$ | $78 \pm 6$ | .006 ($p \sim 0.64$)* |
| Years of education | $8.4 \pm 1.9$ | $6.5 \pm 3.7$ | $-1.99$ ($p < 0.05$)** |
| MMSE | $28.5 \pm 1.6$ | $24 \pm 2.6$ | $-3.6$ ($p < 10^{-3}$)** |

\*Bonferroni and Tukey/Tamhane corrected one-way ANOVA followed by two-independent sample T test if applicable. \*\*Bonferroni and Dunn corrected K samples Kruskal-Wallis test followed by Mann-Whitney U if applicable.

**Table 5.**
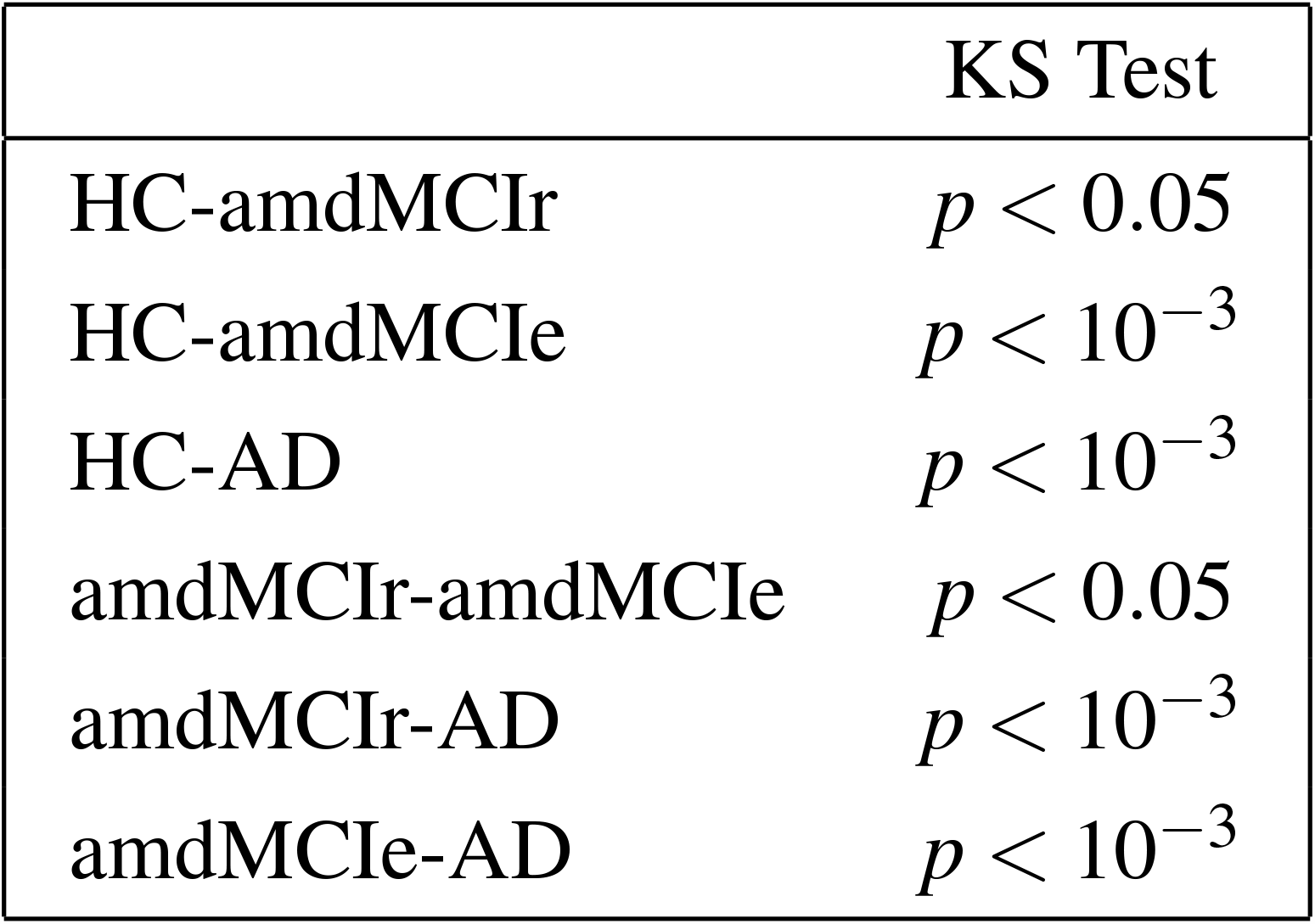
Two sided KS test between groups.

## 3. Discussion

We have characterized the probability density distribution of speech pauses in AD, healthy controls, and two aMCI groups with differential memory impairment profiles, being able to show significant differences across all groups. A gradation has been found in the parameters and shape of the LNDs from AD to HCs, with a-mdMCI-E and a-mdMCI-R standing in the middle of the continuum between those groups (inequations 4 and 5). Moreover, this issue has been addressed in a censoring context affecting shorter pauses which, as it has been shown, lack the reliability of shorter pauses in terms of interrater agreement. For this purpose, we have considered the latest discussions on the fitting of long-tailed distributions [116,119], successfully recovering LNDs for pauses longer than 160*ms*.

Previous discussions addressing the use of these distribution type have proposed temporal thresholds differentiating short pauses from long speech pauses setting this barrier at 268*ms* [94], 323*ms* [120] or more recently at 338*ms* [93]. However, other authors have suggested up to three pause types: short (< 200*ms*), intermediate (200 − 1000*ms*) and long pauses (*>* 1000*ms*) [90]. The classification of “short” and “long” pauses was long reduced to a conceptual discussion about articulatory and/or respiratory (short) pauses on the one hand, and (long) cognitive pauses on the other [121,122]. This theoretical positioning has long gone undisputed-save for some exceptions, see [123,124]-but is currently under question as technical improvements allow for more precise location and measurement of pauses [93,120], and other topics of research has been recently open [125]. We have shown that shorter ranges are affected by higher mean relative errors and lower interrater agreement, which in turn may affect the conclusions drawn from the data (Figure 2). This led us to apply a quantitative method that determines a minimal threshold for pause duration (160*ms*) so that only occurrences beyond that point are considered, a method that can be applied beyond the study of the verbal production in AD. In any case, studying only pauses beyond that threshold may have sense in the context of Alzheimer’s disease as it has been discussed to be related with cognitive generation processes.

Pause duration distribution in speech is adequately explained with a LND that can be inferred with the help of truncated distributions, even when information on shorter ranges is incomplete or censored. To the best of our knowledge, this is the first study where the shape of the lognormally-fitted pause duration distribution is used in the classification of different groups with cognitive impairment in the context of AD. More concretely, the shape and parameters of the LND allow the detection of significant differences in the probability distribution of pauses according to duration across all groups of study (HC, a-mdMCI and AD patients). This pause duration distribution reveals the existence of a continuum from the group with the highest probability of producing short pauses (HC) to the group with the highest probability of making long pauses (AD patients), with a-mdMCI-R performing in the range between HCs and a-mdMCI-E and the latter showing a production pattern more resemblant to that of AD (Figure 4). The confirmation of a higher likelihood to produce more long pauses and less short pauses for those in the group with higher probability of AD progression (a-mdMCI-E) in comparison to the a-mdMCI-R group suggest a new promising tool for dementia prognosis that should be addressed in further studies.

The finding that patients with a-mdMCI-E produce significantly longer pauses with more variance than HC confirms our initial hypothesis and is in line with previous findings [69,59,82]. The gradation found along the AD spectrum and more specifically within the a-mdMCI subtype initially suggests a central role of memory degradation as reflected in the impaired delayed recall and recognition observed in the a-mdMCI-E group in comparison to a-mdMCI-R. Previous studies including correlation analyses of neuropsychological scores and pause duration suggest that longer pause durations arise from difficulties in the retrieval of relevant information from episodic memory both in AD [79] and in MCI due to AD [82], a cognitive domain particularly affected in AD that is compatible with the impaired encoding and consolidation processes observed in our a-mdMCI-E participants.

The fact that other studies using memory-taxing speech tasks failed to find significant differences in mean pause duration [59,68] invites for further enquiry as to criteria for pause labelling and analysis. Our picture description task [106] does not exert the same level of demand on recent anterograde memory while still managing to capture fluency impairments in a-mdMCI patients, in line with previous studies also implementing picture description tasks [59,83,60,64]. In light of this evidence and considering the well documented constellation of semantic and lexical processing deficits in AD [126,127,128,129,130], our results suggest a generalised, more profound deterioration of the memory system from the preclinical stages of Alzheimer’s disease. Our elicitation task successfully taps onto these emerging deficits by imposing a controlled lexicosemantic setting that is also demanding on working memory for discourse building and task maintenance, testing other dimensions of memory in addition to episodic anterograde memory and verbal learning, –which are clearly impaired in these patients– avoiding thus circularity in their diagnosis, which is already based on verbal memory assessment. In MCI the fluency factor is highly correlated with memory measures [60] but to an even greater degree with BNT score (a picture-based test of lexical and semantic memory integrity). However, [64] failed to find such correlations between fluency parameters and psycholinguistic measures in very early MCI. These differences may be the reflection of different stages of the progressive degradation of memory and language that takes place in AD, that in the case of our a-mdMCI-E sample would be at a more advanced phase given their neuropsychological profile and the fact that a considerable number of these individuals progressed to Alzheimer’s dementia within three years (see 1.1. Section). Our two a-mdMCI groups only differed significantly in their memory and pause profile while there were no significant differences in number of months since MCI diagnosis, which suggests that they represent two distinct subtypes with not only differential progression rates, but also eventual outcome and, therefore, prognosis.

While LNDs are very common manifestation in natural sciences, the particular generative process involved in showing this manifestations in speech pause distributions are still not known. Further than being the reminiscence of an unknown multiplicative process, they could be the result of an additive process under some specific constrains [89] or a pattern result of specific neural activity [131]. However, further studies should be carried out to fully understand the production processes and how their deviations are related with healthy disorders. Future studies should also consider the inclusion and comparison of different speech-eliciting tasks in order to clarify the role of memory in the linguistic behaviour of patients in the AD spectrum and evaluate the relative weight of other deficits that might also be at play, in addition to confirming the applicability of this methodology in the design of tests that may serve as early low-cost markers in dementia detection.

## Data Availability

The speech pauses duration corpus used in this study and scripts that ensure reproducibility of all results are public available in https://github.com/ivangtorre/Speech-pause-distribution-as-an-early-marker-for-Alzheimers-disease.
The full complete corpus may be accessible under some considerations upon request to the main author. We have used Python 3.8.2 and R 3.6.3 for the analysis. MLE fit,
Kolmogorov-Smirnov AIC and BIC make use of fitdistrplus 1.1.1 and truncdist 1.0.2.
Numpy, Pandas and Matplotlib libraries are also used. Some other statistical analyses
were performed on IBM SPSS 25.0.
Replicability of the study is addressed through Section 1 (Material and Methods) of the
present paper.

https://github.com/PatriciaPast/Speech-pause-distribution-as-an-early-marker-for-Alzheimers-disease

## 4. Acknowledgements

This work is supported through a PhD grant awarded to P.P-D. by the banking Foundation “La Caixa” (ID 100010434, code: LCF/BQ/ES15/10360020). F.D-V. was supported by a grant awarded to project no. PID2019-107042GB-I00 (Ministerio de Ciencia e Innovación). A.H-F. and I.G.T. were supported by the grant no. TIN2017-89244-R (MACDA) (Ministerio de Economia, Industria y Competitividad, Gobierno de Espanã) and the project PRO2020-S03 (RCO03080 Lingüística Quantitativa) of l’Institut d’Estudis Catalans.

## 6. Reproducibility and replicability

The speech pause duration corpus used in this study and scripts that ensure reproducibility of all results are public available in https://github.com/ivangtorre/Speech-pause-distribution-as-an-early-marker-for-Alzheimers-disease and https://github.com/PatriciaPast/Speech-pause-distribution-as-an-early-marker-for-Alzhei The full complete corpus may be accessible under some considerations upon request to the main author. We have used *Python* 3.8.2 and *R* 3.6.3 for the analysis. MLE fit, Kolmogorov-Smirnov AIC and BIC make use of *fitdistrplus* 1.1.1 and *truncdist* 1.0.2. *Numpy, Pandas* and *Matplotlib* libraries are also used. Some other statistical analyses were performed on *IBM SPSS* 25.0.

Replicability of the study is addressed through Section 1 (*Material and Methods*) of the present paper.

## References

[1] Glynn K, O’Callaghan M, Hannigan O, Bruce I, Gibb M, Coen R, Green E, Lawlor B, Robinson D (2020) Clinical utility of mild cognitive impairment subtypes and number of impaired cognitive domains at predicting progression to dementia: A 20-year retrospective study. International Journal of Geriatric Psychiatry 36, 31–37.

[2] Petersen RC (2018) How early can we diagnose Alzheimer disease (and is it sufficient)? Neurology 91, 395–402.

[3] Livingston G, Sommerlad A, Orgeta V, Costafreda SG, Huntley J, Ames D, Ballard C, Banerjee S, Burns A, Cohen-Mansfield J, Cooper C, Fox N, Gitlin LN, Howard R, Kales HC, Larson EB, Ritchie K, Rockwood K, Sampson EL, Samus Q, Schneider LS, Selbaek G, Teri L, Mukadam N, Cohen-Mansfield J, Gitlin N (2017) Dementia prevention, intervention, and care. Lancet 390, 2673–2734.

[4] Alzheimer’s Association (2018) 2018 Alzheimer’s disease facts and figures. Alzheimer’s Dementia 14, 367–429.

[5] Prince M, Bryce R, Ferri C (2011) World Alzheimer Report 2011: The benefits of early diagnosis and intervention, London.

[6] Callahan BL, Ramirez J, Berezuk C, Duchesne S, Black SE (2015) Predicting Alzheimer’s disease development: A comparison of cognitive criteria and associated neuroimaging biomarkers. Alzheimer’s Res. Ther. 7(1), 68.

[7] Cui Y, Liu B, Luo S, Zhen X, Fan M, Liu T, Zhu W, Park M, Jiang T, Jin JS (2011) Identification of conversion from mild cognitive impairment to alzheimer’s disease using multivariate predictors. PLoS One 6(7), e21896.

[8] Bondi MW, Edmonds EC, Jak AJ, Clark LR, Delano-Wood L,McDonald CR, Nation DA, Libon DJ, A., Galasko D, Salmon DP (2014) Neuropsychological Criteria for Mild Cognitive Impairment Improves Diagnostic Precision, Biomarker Associations, and Progression Rates. Journal of Alzheimer’s Disease 42(1), 275–289.

[9] Jack CR, Bennett DA, Blennow K, Carrillo MC, Dunn B, Haeberlein SB, Holtzman DM, Jagust W, Jessen F, Karlawish J, Liu E, Molinuevo JL, Montine T, Phelps C, Rankin KP, Rowe CC, Scheltens P, Siemers E, Snyder HM, … Silverberg N (2018) NIA-AA Research Framework: Toward a biological definition of Alzheimer’s disease. Alzheimer’s & Dementia 14 (4), 532–562.

[10] Kivipelto M, Mangialasche F, Ngandu T (2018) Lifestyle interventions to prevent cognitive impairment, dementia and Alzheimer disease. Nat. Rev. Neurol. 14, 653–666.

[11] Clark LR, Delano-Wood L, Libon DJ, McDonald CR, Nation DA, Bangen KJ, Jak AJ, Au R, Salmon DP, Bondi MW (2013) Are Empirically-Derived Subtypes of Mild Cognitive Impairment Consistent with Conventional Subtypes? J. Int. Neuropsychol. Soc. 19, 635–645.

[12] Jak AJ, Preis SR, Beiser AS, Seshadri S, Wolf PA, Bondi M W, Au R (2016) Neuropsychological Criteria for Mild Cognitive Impairment and Dementia Risk in the Framingham Heart Study. Journal of the International Neuropsychological Society 22 (9), 937–943.

[13] Petersen RC, Smith GE, Ivnik RJ, Tangalos EG, Schaid DJ, Thibodeau SN, Kokmen E, Waring SC, Kurland LT (1995) Apolipoprotein E Status as a Predictor of the Development of Alzheimer’s Disease in Memory-Impaired Individuals JAMA: The Journal of the American Medical Association 273 (16), 1274–1278.

[14] Petersen RC, Smith GE, Waring SC, Ivnik RJ, Kokmen E, Tangelos EG (1997) Aging, Memory, and Mild Cognitive Impairment. International Psychogeriatric Association 9 (1), 65–69.

[15] Smith GE, Petersen RC, Parisi, JE, Ivnik RJ, Kokmen E, Tangalos EG, Waring S (1996) Definition, course, and outcome of mild cognitive impairment. Aging, Neuropsychology, and Cognition 3 (2), 141–147.

[16] Belleville S, Fouquet C, Hudon C, Zomahoun HTV, Croteau J (2017) Neuropsychological measures that predict progression from mild cognitive impairment to Alzheimer’s type dementia in older adults: a systematic review and meta-analysis. Neuropsychology Rev. 27(4), 328–353.

[17] Fleisher AS, Sowell BB, Taylor C, Gamst AC, Petersen RC, Thal LJ (2007) Clinical predictors of progression to Alzheimer disease in amnestic mild cognitive impairment. Neurology 68 (19), 1588–1595.

[18] Göthlin M, Eckerström M, Rolstad S, Wallin A, Nordlund A (2017) Prognostic Accuracy of Mild Cognitive Impairment Subtypes at Different Cut-Off Levels. Dementia and Geriatric Cognitive Disorders 43, 330–341.

[19] Kim YJ, Cho SK, Kim HJ, Lee JS, Lee J, Jang YK, Vogel JW, Na DL, Kim C, Seo SW (2019) Data-driven prognostic features of cognitive trajectories in patients with amnestic mild cognitive impairments. Alzheimer’s Research and Therapy

[20] Perri R, Serra L, Carlesimo GA, Caltagirone C (2007) Amnestic Mild Cognitive Impairment: Difference of Memory Profile in Subjects Who Converted or Did Not Convert to Alzheimer’s Disease. Neuropsychology 21 (5), 549–558.

[21] Tabert MH, Manly JJ, Liu X, Pelton GH, Rosenblum S, Jacobs M, Zamora D, Goodkind M, Bell K, Stern Y, Devanand DP (2006) Neuropsychological prediction of conversion to alzheimer disease in patients with mild cognitive impairment. Archives of General Psychiatry 63, 916–924.

[22] Aggarwal NT, Wilson RS, Beck TL, Bienias JL, Bennett DA (2005) Mild cognitive impairment in different functional domains and incident Alzheimer’s disease. J. Neurol. Neurosurg. Psychiatry 76, 1479–1484.

[23] Jacobs DM, Sano M, Dooneief G, Marder K, Bell KL, Stern Y (1995) Neuropsychological detection and characterization of preclinical alzheimer’s disease. Neurology 45(5), 957–962.

[24] Grober E, Lipton RB, Hall C, Crystal H (2000) Memory impairment on free and cued selective reminding predicts dementia Neurology 54 827–832.

[25] Mistridis P, Krumm S, Monsch AU, Berres M, Taylor KI (2015) The 12 Years Preceding Mild Cognitive Impairment Due to Alzheimer’s Disease: The Temporal Emergence of Cognitive Decline. Journal of Alzheimer’s Disease 48, 1095–1107.

[26] Chiotis K, Saint-Aubert L, Rodriguez-Vieitez E, Leuzy A, Almkvist O, Savitcheva I, Jonasson M, Lubberink M, Wall A, Antoni G, Nordberg A (2018) Longitudinal changes of tau PET imaging in relation to hypometabolism in prodromal and Alzheimer’s disease dementia. Molecular Psychiatry 23, 1666–1673.

[27] Curiel Cid RE, Crocco EA, Duara R, Garcia JM, Rosselli M, DeKosky ST, Smith G, Bauer R, Chirinos CL, Adjouadi M, Barke W, Loewenstein DA (2020) A novel method of evaluating semantic intrusion errors to distinguish between amyloid positive and negative groups on the Alzheimer’s disease continuum. Journal of Psychiatric Research 124, 131–136.

[28] Goldstein FC, Loring DW, Thomas T, Saleh S, Hajjar I (2019) Recognition Memory Performance as a Cognitive Marker of Prodromal Alzheimer’s Disease. Journal of Alzheimer’s Disease 72, 507–514.

[29] Hessen E, Kirsebom BE, Eriksson CM, Eliassen CF, Nakling AE, Brathen G, Waterloo KK, Aarsland D, Fladby T (2019) In brief neuropsychological assessment, amnestic mild cognitive impairment (MCI) is associated with cerebrospinal fluid biomarkers for cognitive decline in contrast to the prevailing NIA-AA MCI criterion. Journal of Alzheimer’s Disease 67, 715–723.

[30] Putcha D, Brickhouse M, Wolk DA, Dickerson BC (2019) Fractionating the Rey Auditory Verbal Learning Test: Distinct roles of large-scale cortical networks in prodromal Alzheimer’s disease. Neuropsychologia 129, 83–92.

[31] Hampstead BM, Khoshnoodi M, Yan W, Deshpande G, Sathian K (2016) Patterns of effective connectivity during memory encoding and retrieval differ between patients with mild cognitive impairment and healthy older adults. NeuroImage 124, 997–1008.

[32] Prieto Del Val L, Cantero JL, Atienza M (2016) Atrophy of amygdala and abnormal memory-related alpha oscillations over posterior cingulate predict conversion to Alzheimer’s disease. Scientific Reports 6, 31859.

[33] Sarica A, Vasta R, Novellino F, Vaccaro MG, Cerasa A, Quattrone A, for Alzheimer’s Disease Neuroimaging Initiative TADN (2018) MRI Asymmetry Index of Hippocampal Subfields Increases Through the Continuum From the Mild Cognitive Impairment to the Alzheimer’s Disease. Frontiers in Neuro-science

[34] Xie L, Dolui S, Das SR, Stockbower GE, Daffner M, Rao H, Yushkevich PA, Detre JA, Wolk DA (2016) A brain stress test: Cerebral perfusion during memory encoding in mild cognitive impairment. NeuroImage: Clinical 11, 388–397.

[35] Edelman K, Tudorascu D, Agudelo C, Snitz B, Karim H, Cohen A, Mathis C, Price J, Weissfeld L, Klunk W Aizenstein H (2017) Amyloid-β Deposition is Associated with Increased Medial Temporal Lobe Activation during Memory Encoding in the Cognitively Normal Elderly. American Journal of Geriatric Psychiatry 25 (5), 551–560.

[36] Hollands S, Lim YY, Buckley R, Pietrzak RH, Snyder PJ, Ames D, Ellis KA, Harrington K, Lautenschlager N, Martins RN, Masters CL, Villemagne VL, Rowe CC, Maruff P (2014) Amyloid-β Related Memory Decline is not Associated with Subjective or Informant Rated Cognitive Impairment in Healthy Adults. Journal of Alzheimer’s Disease 43 (2), 677–686.

[37] Papp KV, Rentz DM, Mormino EC, Schultz AP, Amariglio RE, Quiroz Y, Johnson KA, Sperling RA (2017) Cued memory decline in biomarker-defined preclinical Alzheimer disease. Neurology 88 (15), 1431–1438.

[38] Hanseeuw BJ, Betensky RA, Jacobs HIL, Schultz AP, Sepulcre J, Becker JA, Cosio DMO, Farrell M, Quiroz YT, Mormino EC, Buckley RF, Papp KV, Amariglio RA, Dewachter I, Ivanoiu A, Huijbers W, Hedden T, Marshall GA, Chhatwal JP, … Johnson K (2019) Association of Amyloid and Tau With Cognition in Preclinical Alzheimer Disease. JAMA Neurology 76 (8), 915–924.

[39] Marks SM, Lockhart SN, Baker SL, Jagust WJ (2017) Tau and B-amyloid are associated with medial temporal lobe structure, function, and memory encoding in normal aging. Journal of Neuroscience 37 (12), 3192–3201.

[40] Sperling RA, Mormino EC, Schultz AP, Betensky RA, Papp KV, Amariglio RE, Hanseeuw BJ, Buckley R, Chhatwal J, Hedden T, Marshall GA, Quiroz YT, Donovan NJ, Jackson J, Gatchel JR, Rabin JS, Jacobs H, Yang HS, Properzi M, … Johnson KA (2019) The impact of amyloid-β and tau on prospective cognitive decline in older individuals. Annals of Neurology 85 (2), 181–193.

[41] Tort-Merino A, Olives J, León M, Penãloza C, Valech N, Santos-Santos MA, Càmara E, Grönholm-Nyman P, Martínez-Lage P, Fortea J, Molinuevo JL, Sánchez-Valle R, Laine M, Rodríguez-Fornells A, Rami L (2019) Tau Protein is Associated with Longitudinal Memory Decline in Cognitively Healthy Subjects with Normal Alzheimer’s Disease Cerebrospinal Fluid Biomarker Levels. Journal of Alzheimer’s Disease 70, 211–225.

[42] Gagliardi G, Epelbaum S, Houot M, Bakardjian H, Boukadida L, Revillon M, Dubois B, Dalla Barba G, La Corte V (2019) Which Episodic Memory Performance is Associated with Alzheimer’s Disease Biomarkers in Elderly Cognitive Complainers? Evidence from a Longitudinal Observational Study with Four Episodic Memory Tests (Insight-PreAD). Journal of Alzheimer’s Disease 70, 811–824.

[43] Ochoa JF, Alonso JF, Duque JE, Tobón CA, Manãnas MA, Lopera F, Hernández AM (2017) Successful object encoding induces increased directed connectivity in presymptomatic early-onset Alzheimer’s disease. Journal of Alzheimer’s Disease 55, 1195–1205.

[44] Rabipour S, Rajagopal S, Yu E, Pasvanis S, Lafaille-Magnan ME, Breitner J, Rajah MN (2020) APOE4 Status is Related to Differences in Memory-Related Brain Function in Asymptomatic Older Adults with Family History of Alzheimer’s Disease: Baseline Analysis of the PREVENT-AD Task Functional MRI Dataset. Journal of Alzheimer’s Disease 76, 97–119.

[45] Karr JE, Graham RB, Hofer SM, Muniz-Terrera G (2018) When does cognitive decline begin? A systematic review of change point studies on accelerated decline in cognitive and neurological outcomes preceding mild cognitive impairment, dementia, and death. Psychology and Aging 33 (2), 195–218.

[46] Mura T, Proust-Lima C, Jacqmin-Gadda H, Akbaraly TN, Touchon J, Dubois B, Berr C (2014) Measuring cognitive change in subjects with prodromal Alzheimer’s disease. J. Neurol. Neurosurg. Psychiatry 85(4), 363–370.

[47] Belleville S, Gauthier S, Lepage É, Kergoat MJ, Gilbert B (2014) Predicting decline in mild cognitive impairment: A prospective cognitive study. Neuropsychology 28(4), 643.

[48] Cloutier S, Chertkow H, Kergoat MJ, Gauthier S, Belleville, S (2015) Patterns of cognitive decline prior to dementia in persons with mild cognitive impairment.J Alzheimers Dis 47(4), 901–913.

[49] Pravatà E, Tavernier J, Parker R, Vavro H, Mintzer JE, Spampinato MV (2016) The neural correlates of anomia in the conversion from mild cognitive impairment to Alzheimer’s disease. Neuroradiology 58(1), 59–67.

[50] Kim BS, Kim YB, Kim, H (2019) Discourse measures to differentiate between mild cognitive impairment and healthy aging. Frontiers in Aging Neuroscience 11, 221.

[51] Mazzeo S, Santangelo R, Bernasconi MP, Cecchetti G, Fiorino A, Pinto P, Magnani, G (2016). Combining cerebrospinal fluid biomarkers and neuropsychological assessment: a simple and cost-effective algorithm to predict the progression from mild cognitive impairment to Alzheimer’s disease dementia. J Alzheimers Dis 54(4), 1495–1508.

[52] Vaughan RM, Coen RF, Kenny R, Lawlor BA (2018) Semantic and Phonemic Verbal Fluency Discrepancy in Mild Cognitive Impairment: Potential Predictor of Progression to Alzheimer’s Disease. J. Am. Geriatr. Soc. 66(4), 755–759.

[53] Vita MG, Marra C, Spinelli P, Caprara A, Scaricamazza E, Castelli D, Quaranta D (2014) Typicality of Words Produced on a Semantic Fluency Task in Amnesic Mild Cognitive Impairment: Linguistic Analysis and Risk of Conversion to Dementia. J Alzheimers Dis 42, 1171–1178.

[54] Gainotti G, Quaranta D, Vita MG, Marra C (2014) Neuropsychological predictors of conversion from mild cognitive impairment to Alzheimer’s disease. J Alzheimers Dis 38(3), 481–495.

[55] Gleason CE, Norton D, Anderson ED, Wahoske M, Washington DT, Umucu E, … Asthana S. (2018) Cognitive variability predicts incident Alzheimer’s disease and mild cognitive impairment comparable to a cerebrospinal fluid biomarker. J Alzheimers Dis 61(1), 79–89.

[56] Filiou RP, Bier N, Slegers A, Houzé B, Belchior P, Brambati SM (2020). Connected speech assessment in the early detection of Alzheimer’s disease and mild cognitive impairment: a scoping review. Aphasiology, 34(6), 723–755.

[57] Mueller KD, Hermann B, Mecollari J, Turkstra LS (2018) Connected speech and language in mild cognitive impairment and Alzheimer’s disease: A review of picture description tasks. J. Clin. Exp. Neuropsychol. 40(9), 917–939.

[58] Asgari M, Kaye J, Dodge H (2017) Predicting mild cognitive impairment from spontaneous spoken utterances. Alzheimer’s and Dementia: Translational Research and Clinical Interventions 3, 219–228.

[59] Beltrami D, Gagliardi G, Favretti RR, Ghidoni E, Tamburini F, Calzà L (2018) Speech Analysis by Natural Language Processing Techniques: A Possible Tool for Very Early Detection of Cognitive Decline? Front. Aging Neurosci. 10, 369.

[60] Fraser KC, Lundholm Fors K, Eckerström M.Öhman F, Kokkinakis D (2019) Predicting MCI Status From Multimodal Language Data Using Cascaded Classifiers. Front, Aging Neurosci. 11

[61] Gosztolya G, Vincze V, Tóth L, Pákáski M, Kálmán J, Hoffmann I (2019) Identifying Mild Cognitive Impairment and mild Alzheimer’s disease based on spontaneous speech using ASR and linguistic features. Computer Speech and Language 53, 181–197.

[62] König A, Satt A, Sorin A, Hoory R, Toledo-Ronen O, Derreumaux A, Manera V, Verhey F, Aalten P, Robert PH, David R (2015) Automatic speech analysis for the assessment of patients with predementia and Alzheimer’s disease. Alzheimer’s Dement. Diagnosis, Assess. Dis. Monit. 1, 112–124.

[63] Mirheidari B, Blackburn D, Walker T, Reuber M, Christensen H (2019) Dementia detection using automatic analysis of conversations. Computer Speech and Language 53, 65–79.

[64] Mueller KD, Koscik RL, Hermann BP, Johnson SC, Turkstra LS (2018) Declines in connected language are associated with very early mild cognitive impairment: Results from the Wisconsin Registry for Alzheimer’s Prevention. Front. Aging Neurosci.

[65] Orimaye SO, Wong JSM, Golden KJ, Wong CP, Soyiri IN (2017) Predicting probable Alzheimer’s disease using linguistic deficits and biomarkers. BMC Bioinformatics

[66] Orimaye SO, Wong JSM, Wong CP (2018) Deep language space neural network for classifying mild cognitive impairment and Alzheimer-type dementia. PLOS ONE 13(11), e0205636.

[67] Rentoumi V, Raoufian L, Ahmed S, De Jager CA, Garrard P (2014) Features and machine learning classification of connected speech samples from patients with autopsy proven Alzheimer’s disease with and without additional vascular pathology. J Alzheimers Dis 42, S3–S17.

[68] Roark B, Mitchell M, Hosom JP, Hollingshead K, Kaye J (2011) Spoken language derived measures for detecting mild cognitive impairment. IEEE Trans. Audio, Speech Lang. Process. 19, 2081–2090.

[69] Toth L, Hoffmann I, Gosztolya G, Vincze V, Szatloczki G, Banreti Z, Pakaski M, Kalman J (2018) A Speech Recognition-based Solution for the Automatic Detection of Mild Cognitive Impairment from Spontaneous Speech. Current Alzheimer’s Research 15, 130–138.

[70] Lopez-de-Ipina K, Martinez-de-Lizarduy U, Calvo PM, Beitia B, Garcia-Melero J, Ecay-Torres M, Estanga A, Faundez-Zanuy M (2017) Analysis of Disfluencies for automatic detection of Mild Cognitive Impartment: a deep learning approach. 2017 International Conference and Workshop on Bioinspired Intelligence (IWOBI) Funchal, 2017, 1–4.

[71] Meilán JJG, Martínez-Sánchez F, Carro J, Carcavilla N, Ivanova O (2018) Voice Markers of Lexical Access in Mild Cognitive Impairment and Alzheimer’s Disease. Current Alzheimer Research 15, 111–119.

[72] Themistocleous C. Eckerström M, Kokkinakis D (2020) Voice quality and speech fluency distinguish individuals with Mild Cognitive Impairment from Healthy Controls. PLoS ONE

[73] Abdalla M, Rudzicz F, Hirst G (2018) Rhetorical structure and Alzheimer’s disease. Aphasiology 32 (1), 41–60.

[74] Drummond C, Coutinho G, Fonseca RP, Assunção N, Teldesch, A, de Oliveira-Souza R, Moll J, TovarMoll F, Mattos P (2015) Deficits in narrative discourse elicited by visual stimuli are already present in patients with mild cognitive impairment. Front. Aging Neurosci.

[75] Pompili A, Abad A, Martins de Matos D, Pavao Martins I (2018) Topic coherence analysis for the classification of Alzheimer’s disease. IberSPEECH 2018, 281–285.

[76] Toledo CM, Aluisio SM, Dos Santos LB, Brucki SMD, Tres ES, de Oliveira MO, Mansur LL (2018) Analysis of macrolinguistic aspects of narratives from individuals with Alzheimer’s disease, mild cognitive impairment, and no cognitive impairment. Alzheimer’s & Dementia 10, 31–40.

[77] Gayraud F, Lee HR, Barkat-Defradas M (2011) Syntactic and lexical context of pauses and hesitations in the discourse of Alzheimer patients and healthy elderly subjects. Clinical linguistics & phonetics 25(3), 198–209.

[78] Hoffmann I, Nemeth D, Dye CD, Pákáski M, Irinyi T, Kálmán J (2010) Temporal parameters of spontaneous speech in Alzheimer’s disease. International Journal of Speech-Language Pathology 12(1), 29–34.

[79] Pistono A, Pariente J, Bézy C, Lemesle B, Le Men J, Jucla M (2019) What happens when nothing happens? An investigation of pauses as a compensatory mechanism in early Alzheimer’s disease. Neuropsychologia 124, 133–143.

[80] Singh S, Bucks RS, Cuerden JM (2001) Evaluation of an objective technique for analysing temporal variables in DAT spontaneous speech. Aphasiology 15(6), 571–583.

[81] Fraser KC, Meltzer JA, Rudzicz F (2016) Linguistic features identify Alzheimer’s disease in narrative speech. J Alzheimers Dis 49(2), 407–422.

[82] Pistono A, Jucla M, Barbeau EJ, Saint-Aubert L, Lemesle B, Calvet B, Köpke B, Puel M, Pariente J (2016) Pauses during Autobiographical Discourse Reflect Episodic Memory Processes in Early Alzheimer’s Disease. J Alzheimers Dis 50, 687–698.

[83] Sluis RA, Angus D, Wiles J, Back A, Gibson TA, Liddle J, Worthy P, Copland D, Angwin AJ (2020) An Automated Approach to Examining Pausing in the Speech of People With Dementia. Am. J. Alzheimers. Dis. Other Demen. 35, 1533317520939773.

[84] Goldman-Eisler, F. (1961). The distribution of pause durations in speech. Language and Speech, 4(4), 232–237.

[85] Chien, J. T., & Huang, C. H. (2003). Bayesian learning of speech duration models. IEEE Transactions on Speech and Audio Processing, 11(6), 558–567.

[86] Rosen KM, Kent RD, Duffy JR (2003) Lognormal distribution of pause length in ataxic dysarthria. Clinical Linguistics & Phonetics 17, 469–486.

[87] Rosen, K. M. (2005). Analysis of speech segment duration with the lognormal distribution: A basis for unification and comparison. Journal of Phonetics, 33(4), 411–426.

[88] Torre, I. G., Luque, B., Lacasa, L., Luque, J., & Hernández-Fernández, A. (2017). Emergence of linguistic laws in human voice. Scientific reports, 7, 43862.

[89] Torre, I. G., Luque, B., Lacasa, L., Kello, C. T., & Hernández-Fernández, A. (2019). On the physical origin of linguistic laws and lognormality in speech. Royal Soc Op Sci 6(8), 191023.

[90] Campione E, Véronis J (2002) A large-scale multilingual study of silent pause duration. In Proceedings of Speech Prosody 2002, 199–202.

[91] Goldman J-P, Thomas F, Roekhaut S, Simon A-C (2010) É tude Statistique De La Durée Pausale Dans Différents Styles De Parole. Journées d’Etudes sur la Parol. In: Actes des 28èmes journées d’étude sur la parole (JEP), 2010, 161–164.

[92] Bailly G, Gouvernayre C (2012) Pauses and respiratory markers of the structure of book reading. In 13th Annual Conference of the International Speech Communication Association 2012, INTERSPEECH 2012, 2218–2221.

[93] Angelopoulou G, Kasselimis D, Makrydakis G, Varkanitsa M, Roussos P, Goutsos D, Evdokimidis I, Potagas C (2018) Silent pauses in aphasia. Neuropsychologia 114, 41–49.

[94] Hird K, Kirsner K (2010) Objective measurement of fluency in natural language production: A dynamic systems approach. J. Neurolinguistics 23(5), 518–530.

[95] McKhann G, McKhann G, Drachman D, Drachman D, Folstein M, Folstein M, Katzman R, Katzman R, Price D, Price D, Stadlan EM, Stadlan EM (1984) Clinical diagnosis of Alzheimer’s disease: report of the NINCDS-ADRDA Work Group under the auspices of Department of Health and Human Services Task Force on Alzheimer’s Disease. Neurology 34, 939–944

[96] McKhann GM, Knopman DS, Chertkow H, Hyman BT, Jack CR, Kawas CH, Klunk WE, Koroshetz WJ, Manly JJ, Mayeux R, Mohs RC, Morris JC, Rossor MN, Scheltens P, Carrillo MC, Thies B, Weintraub S, Phelps CH (2011) The diagnosis of dementia due to Alzheimer’s disease: Recommendations from the National Institute on Aging-Alzheimer’s Association workgroups on diagnostic guidelines for Alzheimer’s disease. Alzheimer’s Dementia 7(3), 263–269.

[97] Hughes CP, Berg L, Danziger WL (1982) A new clinical scale for the staging of dementia. Br. J. Psychiatry 140, 566–572.

[98] Petersen RC (2004) Mild cognitive impairment as a clinical entity and treatment target. Arch. Neurol. 62, 1160–1163; discussion 1167.

[99] Albert MS, Dekosky ST, Dickson D, Dubois B, Feldman HH, Fox NC, Gamst A, Holtzman DM, Jagust WJ, Petersen RC, Snyder PJ, Carrillo MC, Thies B, Phelps CH (2011) The diagnosis of mild cognitive impairment due to Alzheimer’s disease: Recommendations from the National Institute on Aging-Alzheimer’s Association workgroups on diagnostic guidelines for Alzheimer’s disease. Alzheimer’s Dementia 7, 270–279.

[100] Rey A (1964) L’examen clinique en Psychologie, Presses Universitaires de France, Paris.

[101] Folstein MF, Folstein SE, McHugh PR (1975) “Mini-mental state”. A practical method for grading the cognitive state of patients for the clinician. J. Psychiatr. Res. 12, 189–198.

[102] Kaplan E, Goodglass H WS (1983) The Boston Naming Test, Lea and Febiger, Philadelphia.

[103] Wechsler D (1997) WAIS-III: Wechsler Adult Intelligence Scale (3rd ed.) Administration and scoring manual, The Psychological Corporation, San Antonio.

[104] Sunderland T, Hill JL, Mellow AM, Lawlor BA, Gundersheimer J, Newhouse PA, Grafman JH (1989) Clock Drawing in Alzheimer’s Disease: A Novel Measure of Dementia Severity. J. Am. Geriatr. Soc. 37, 725729.

[105] Della Sala S, Laiacona M, Trivelli C, Spinnler H (1996) Poppelreuter-Ghent’s overlapping figures test: its sensitivity to age, and its clinical use. Arch. Clin. Neuropsychol. 10, 511–534.

[106] Paradis M (1987) Bilingual aphasia test, Lawrence Erlbaum Associates, Inc., Hillsdale, NJ.

[107] Boersma P, Weenink D (2007) Praat: doing phonetics by computer (Version 4.5.)[Computer program]. Retrieved from http://www.praat.org/ 5, 341–345.

[108] López-de-Ipinã K, Martinez-de-Lizarduy U, Calvo PM, Beitia B, García-Melero J, Fernández E, Ecay-Torres M, Faundez-Zanuy M, Sanz P (2018) On the analysis of speech and disfluencies for automatic detection of Mild Cognitive Impairment. Neural Comput. Appl. 1–9.

[109] Buchanan TW, Laures-Gore JS, Duff MC (2014) Acute stress reduces speech fluency. Biol. Psychol. 97, 60–66.

[110] Verfaillie SCJ, Witteman J, Slot RER, Pruis IJ, Vermaat LEW, Prins ND, Schiller NO, van de Wiel M, Scheltens P, van Berckel BNM, van der Flier WM, Sikkes SAM (2019) High amyloid burden is associated with fewer specific words during spontaneous speech in individuals with subjective cognitive decline. Neuropsychologia 131, 184–192.

[111] Fraundorf SH, Watson DG (2013) Alice’s adventures in um-derland: Psycholinguistic sources of variation in disfluency production. Lang Cogn Process 29, 1083–1096.

[112] Clark HH, Fox Tree JE (2002) Using uh and um in spontaneous speaking. Cognition 84, 73–111.

[113] Pakhomov SVS, Smith GE, Chacon D, Feliciano Y, Graff-Radford N, Caselli R, Knopman DS (2010) Computerized analysis of speech and language to identify psycholinguistic correlates of frontotemporal lobar degeneration. Cogn. Behav. Neurol. 23, 165–177.

[114] Crow EL, Shimizu K. (1987) Lognormal distributions. New York: Marcel Dekker.

[115] Hernández-Fernández A, G Torre I, Garrido JM, Lacasa L (2019) Linguistic laws in speech: the case of Catalan and Spanish. Entropy 21(12), 1153.

[116] Clauset A, Shalizi CR, Newman ME (2009) Power-law distributions in empirical data.SIAM review 51(4), 661–703.

[117] Eliason SR (1993) Maximum likelihood estimation: Logic and practice (No. 96). Sage, Newbury Park, CA.

[118] Pawitan Y (2001) In all likelihood: statistical modelling and inference using likelihood. Oxford University Press.

[119] Corral Á, González Á (2019) Power law size distributions in geoscience revisited. Earth and Space Sci. 6(5), 673–697.

[120] Rosen K, Murdoch B, Folker J, Vogel A, Cahill L, Delatycki M, Corben L (2010) Automatic method of pause measurement for normal and dysarthric speech. Clinical Linguistics & Phonetics 24(2), 141–154.

[121] Goldman-Eisler F (1972) Pauses, clauses, sentences. Lang. and speech 15(2), 103–113.

[122] Dalton P, Hardcastle WJ (1989) Disorders of fluency. New York: Wiley-Blackwell.

[123] Hieke AE, Kowal S, O’Connell DC (1983) The trouble with “Articulatory” pauses. Language and Speech 26(3), 203–214.

[124] Picheny MA, Durlach NI, Braida LD (1986) Speaking clearly for the hard of hearing II: Acoustic characteristics of clear and conversational speech. Speech, Lang and Hear Res 29(4), 434–446.

[125] Krivokapic J, Styler W, Parrell B (2020) Pause postures: The relationship between articulation and cognitive processes during pauses. Journal of Phonetics 79, 100953.

[126] Duong A, Whitehead V, Hanratty K, Chertkow H (2006) The nature of lexico-semantic processing deficits in mild cognitive impairment. Neuropsychologia 44 (10), 1928–1935.

[127] Gardini S, Venneri A, Sambataro F, Cuetos F, Fasano F, Marchi M, Crisi G, Caffarra P (2015) Increased Functional Connectivity in the Default Mode Network in Mild Cognitive Impairment: A Maladaptive Compensatory Mechanism Associated with Poor Semantic Memory Performance. J Alzheimers Dis 45 (2), 457–470.

[128] Joubert S, Felician O, Barbeau EJ, Didic M, Poncet M, Ceccaldi M (2008) Patterns of semantic memory impairment in Mild Cognitive Impairment. Behavioural Neurology 19, 35–40.

[129] Pineault J, Jolicoeur P, Grimault S, Bermudez P, Brambati SM, Lacombe J, Villalpando JM, Kergoat MJ, Joubert S (2018) Functional changes in the cortical semantic network in amnestic mild cognitive impairment. Neuropsychology 32 (4), 417–435.

[130] Taler V, Monetta L, Sheppard C, Ohman A (2019) Semantic Function in Mild Cognitive Impairment. Frontiers in Psychology

[131] Buzsáki G, Mizuseki K (2014) The log-dynamic brain: how skewed distributions affect network operations. Nat. Rev. Neurosci. 15, 264–278.

